# Genomic Insights into Pediatric Intestinal Inflammatory and Eosinophilic Disorders using Single-sell RNA-sequencing

**DOI:** 10.1101/2023.09.26.23295909

**Authors:** Marissa R. Keever-Keigher, Lisa Harvey, Veronica Williams, Carrie A. Vyhlidal, Atif A. Ahmed, Jeffery J. Johnston, Daniel A. Louiselle, Elin Grundberg, Tomi Pastinen, Craig A. Friesen, Rachel Chevalier, Craig Smail, Valentina Shakhnovich

**Author notes:** **Correspondence:** Craig Smail. Joint senior authors (Craig Smail, Valentina Shakhnovich). with Ironwood Pharmaceuticals has no scientific or financial bearing on the work presented herein (Valentina Shakhnovich).

## Abstract

Chronic inflammation of the gastrointestinal tissues underlies gastrointestinal inflammatory disorders, leading to tissue damage and a constellation of painful and debilitating symptoms. These disorders include inflammatory bowel diseases (Crohn’s disease and ulcerative colitis), and eosinophilic disorders (eosinophilic esophagitis and eosinophilic duodenitis). Gastrointestinal inflammatory disorders can often present with overlapping symptoms necessitating the use of invasive procedures to give an accurate diagnosis. This study used peripheral blood mononuclear cells from individuals with Crohn’s disease, ulcerative colitis, eosinophilic esophagitis, and eosinophilic duodenitis to better understand the alterations to the transcriptome of individuals with these diseases and identify potential markers of active inflammation within the peripheral blood of patients that may be useful in diagnosis. Single-cell RNA-sequencing was performed on peripheral blood mononuclear cells isolated from the blood samples of pediatric patients diagnosed with gastrointestinal disorders, including Crohn’s disease, ulcerative colitis, eosinophilic esophagitis, eosinophilic duodenitis, and controls with histologically healthy gastrointestinal tracts. We identified 730 (FDR < 0.05) differentially expressed genes between individuals with gastrointestinal disorders and controls across eight immune cell types. There were common patterns among GI disorders, such as the widespread upregulation of *MTRNR2L8* across cell types, and many differentially expressed genes showed distinct patterns of dysregulation among the different gastrointestinal diseases compared to controls, including upregulation of *XIST* across cell types among individuals with ulcerative colitis and upregulation of Th2-associated genes in eosinophilic disorders. These findings indicate both overlapping and distinct alterations to the transcriptome of individuals with gastrointestinal disorders compared to controls, which provide insight as to which genes may be useful as markers for disease in the peripheral blood of patients.

## Introduction

In genetically predisposed individuals, chronic overactivation of the inflammatory response damages tissues along the gastrointestinal (GI) tract frequently resulting in painful and debilitating symptoms (1, 2). GI inflammatory disorders include inflammatory bowel disease (IBD) and eosinophilic gastrointestinal diseases such as eosinophilic esophagitis (EoE) and eosinophilic duodenitis (EoD). IBD is characterized by chronic relapsing neutrophilic inflammation of the intestine and can be divided into two main subtypes based on the site and characteristics of inflammation, with Crohn’s disease (CD) occurring within any portion of the gut and ulcerative colitis (UC) being confined to the colon. IBD affects patients of all ages, with approximately a quarter of patients are diagnosed before adulthood and incidence of pediatric IBD is increasing (3, 4). EoE and EoD are Th-2 mediated inflammatory disorders which can cause dysphagia, vomiting, abdominal pain, and stricturing. Histologically EoE and EoD are characterized by mucosal eosinophilia (5) and may exist independently or as a comorbid condition with either form of IBD. However eosinophilic infiltration of the mucosa may also precede histologic evidence of IBD (crypt distortion, cryptitis with crypt abscesses, mucus depletion from goblet cells, granulomas), sometimes by years (6, 7), further complicating the ability to differentiate concomitant eosinophilic disease from early harbingers of IBD.

Differentiating between UC and CD or between early IBD and EoE/EoD can help direct therapy. As example, CD patients benefit from early biologic therapy (8), and colectomy is only curative in UC (9). Patients with two concomitant diseases (i.e. EoE and CD) may affect the same area but exhibit different histology and symptoms and require different treatments (10). Additionally, early knowledge of whether mucosal eosinophilia is burgeoning IBD can allow early and appropriate intervention. The pediatric population with IBD and EoE/EoD are particularly vulnerable to growth failure (11, 12) and interruptions in social-emotional development (13) and will require decades of healthcare for their condition. Since disease diagnosis and follow up evaluation currently require invasive endoscopic testing, non-invasive diagnostics and targeted therapies are particularly valuable to this subset of patients.

Characterizing the role of specific immune cell populations in GI diseases has aided in recognizing aberrant processes that underlie these conditions, initiating an important shift in the treatment paradigm away from systemic, non-targeted immunosuppression (fraught with many unwanted side effects) to targeted modulation at the site of disease activity (14). Continued identification of novel therapeutic targets and molecular signatures of disease is pivotal for advancing and optimizing treatment options for chronic immune-mediated inflammatory disorders. In the IBD-affected GI tract, dendritic cells (DCs)—antigen presenting cells belonging to the innate immune system—exhibit up-regulation of microbial recognition receptors and increased cytokine production (15) that appears to induce inflammation through activation of T cells (16). T cells play a crucial role in immune homeostasis (17, 18), and dysregulation of cytokine signaling in CD4^+^ T cells of the GI tract has been shown to lead to pathogenic inflammation (18, 19). T cells also play a key role in eosinophilic disorders of the GI tract, as overexpression of interleukin 5 (IL-5) in CD2^+^ T cells is sufficient to produce eosinophilia in the esophagus and small intestine of transgenic mice (20).

In addition to contributing to inflammation and tissue damage at lesion sites in the GI tract, evidence of altered gene expression and signaling among immune cells in peripheral blood may be reflective of luminal inflammation (21-23). Information gathered from peripheral blood has the potential to identify minimally-invasive, diagnosis-specific and/or disease location-specific genetic markers for GI diseases. Discovery of such biomarkers could potentially decrease the need for repeat endoscopy, which is invasive, associated with risks, and costly. Furthermore, identification of altered gene expression within these disorders at the cellular level could yield a more complete understanding of impacted pathways within specific cell types, and aid in characterizing genetic signatures for future use in disease sub-typing and drug response applications. However, due to the complex and multifactorial nature of GI diseases and differences in immune cell response across GI disease sub-types, reliable indicators of active inflammation have been difficult to characterize within the peripheral blood of patients to date.

In this study, we identified cell-type specific differential gene expression and enrichment of functional gene ontology terms and pathways in individuals diagnosed with CD, UC, EoE, and EoD using single-cell RNA-sequencing of peripheral blood mononuclear cells (PBMCs) in a pediatric patient cohort. Results from this study assist in uncovering the genomic landscape of these phenotypes, which often present with overlapping symptoms in patients, and aid in identifying robust markers of disease types within the peripheral blood mononuclear cells of patients.

## Materials and Methods

### Patient Information

Potential study participants were identified via review of the clinical endoscopy schedule and the electronic medical record (EMR) at the Children’s Mercy Hospital (CMH), a tertiary regional pediatric hospital in the Midwestern United States. To be considered for study inclusion, patients had to be between 1 month and 21 years of age (inclusive), undergoing both upper and lower endoscopy with biopsies for clinical purposes, having a reasonable clinical suspicion for a new diagnosis of immune-mediated inflammatory disease or another clinical indication for undergoing endoscopy (e.g., abdominal pain), and not receiving systemic immunomodulating, immunosuppressive, or biological drugs. Subjects were recruited on the day of procedure, prior to endoscopy. All subjects were fasting at least 8 hours for procedural purposes as part of routine medical care. Only those subjects who provided informed consent (if 18 years of age), or informed assent with parental/legal guardian consent (if under 18 years of age) were included. All research activities were approved by the CMH Institutional Review Board. A total of 35 patients seen in the CMH operating room for routine endoscopy (Kansas City, MO, USA) were included in the study. This cohort consists of 16 males and 19 females ranging in age from 6.17 to 19.25 years with a mean age of 13.3 years. Seven patients were subsequently diagnosed with CD, nine with EoD, ten with EoE, and three with UC. Six patients were identified as controls who had no relevant GI pathology on visual or histologic examination of tissue. Review of individuals’ medical charts indicated no bias toward a single drug therapy in any sub-cohort.

### PBMC Isolation

Up to 4 mL of whole blood was collected from patients in a sodium heparin tube and stored on ice until PBMCs were isolated. Automated PBMC isolation was performed using a STEMCELL Technologies RoboSep-S using the EasySep Direct Human PBMC Isolation Kit (STEMCELL Technologies Cat No. 19654RF) and following the manufacturer’s protocol. After PBMC isolation, the resulting cell suspension was centrifuged at 300 x g for 8 min, and the supernatant was carefully aspirated. The cell pellet was resuspended in 1 mL of ACK Lysing Buffer (Thermo Fisher Cat No. A1049201) and incubated at room temperature for 5 min to remove any remaining RBCs. The cell suspension was centrifuged at 300 x g for 8 min, and the supernatant was carefully aspirated. Cells were washed twice with PBS (Thermo Fisher Cat No. 14190144) supplemented with 2% heat-inactivated FBS (GE Healthcare Cat No. SH30088.03HI), and cell count and viability were assessed using a Countess II automated cell counter. An aliquot of 300,000 cells was diluted in a total volume of 200 μL of PBS + 2% FBS and frozen at -80°C for downstream DNA isolation and genotyping.

The remaining cells were cryopreserved in aliquots of at least one million cells by centrifuging at 300 x g for 8 min, aspirating the supernatant, and resuspending the cell pellets in Recovery Cell Culture Freezing Medium (Thermo Fisher Cat No. 12648010). The cell suspensions were transferred to cryogenic storage vials and were slow-frozen overnight to a temperature of -80°C in a Corning CoolCell FTS30.

### DNA Isolation and Genotyping

Aliquots of 300,000 PBMCs frozen in PBS + 2% FBS were thawed at room temperature, and DNA was isolated using the Qiagen DNeasy Blood & Tissue Kit (Qiagen Cat No. 69506) according to the manufacturer’s protocol. Eluate was concentrated to approximately 50 μL using an Eppendorf Vacufuge Plus, and DNA was quantified using a Qubit dsDNA BR Assay Kit following the manufacturer’s protocol. All DNA samples were selected for high-density genotyping using the Illumina Global Screening Array (GSAMD-24v1-0) according to protocols recommended by Illumina.

### Cell Pooling

Two pools of PBMCs were made. Thawing Medium for PBMC samples consisted of IMDM (ATCC Cat No. 30-2005) supplemented with 10% heat-inactivated FBS, 100 units/mL of penicillin, and 100 μg/mL of streptomycin. For each sample to be thawed, 10 mL of Thawing Medium was prewarmed in a 37°C bead bath. Cells were thawed in groups of up to five samples at a time. The cryovials were placed in a 37°C bead bath. When thawed, the cryovials and 15-mL conical tubes containing Thawing Medium were aseptically transferred to the biosafety cabinet. For each sample, 1 mL of Thawing Medium was added, dropwise, to the cell suspension. The cell suspension was pipette-mixed and then diluted in the remaining 9 mL of Thawing Medium. The thawed and diluted cells were left at room temperature while the remaining cells to be pooled were similarly thawed. When all samples were thawed, the samples were centrifuged at 300 x g for 8 min. The supernatant was carefully aspirated, and the cell pellets were resuspended in 0.5 mL of room-temperature Thawing Medium. All samples were placed on ice and then pooled together. The pool was passed through a 40-μm nylon mesh cell strainer to remove cell aggregates. The pool was centrifuged at 300 x g for 8 min at 4°C, and the supernatant was carefully aspirated. The cell pellet was resuspended in 1 mL of cold Thawing Medium, and cell count and viability were assessed using a Countess II automated cell counter. No fewer than three aliquots per pool were cryopreserved by centrifuging at 300 x g for 8 min at 4°C and resuspending the cell pellets in Recovery Cell Culture Freezing Medium. The cell suspensions were transferred to cryogenic storage vials and were slow-frozen overnight to a temperature of -80°C in a Corning CoolCell FTS30.

### Single-cell Sequencing

Aliquots from each pool were thawed for scRNA-seq. The 10x Genomics Chromium Single Cell 3LJ Reagent Kit v3 was used according to the manufacturer’s protocol to target approximately 15,000 cells per scRNA-seq capture. Libraries were sequenced on an Illumina NovaSeq 6000 platform using 2x94 cycle paired-end sequencing.

### scRNA-seq Alignment and Quality Control

CellRanger v 4.0.0 (10x Genomics) was used for read alignment to the GRCh38 (2020) reference genome, gene counting, and cell calling. Demuxlet (24) was used to demultiplex single-cell data, assigning reads back to the patient of origin using VCF files associated with each patient. Additionally, deumuxlet was used to remove data in instances where barcodes were assigned to more than a single cell.

Quality control of cells was performed with the Seurat v 4.9.9 (25) package in R v 4.2.1. Cells with greater than 20% MT-RNA, fewer than 500 UMIs, and fewer than 0.8 log_10_ genes per UMI were removed. Annotation of the cell types of remaining cells was performed with the Azimuth v 0.4.6 package in R using a previously published PBMC reference (26) to annotate cells to eight broad level one cell types and 30 more specific level two cell types (**Supplementary File 1 Table A**).

### Pseudobulk Differential Expression Analysis and Functional Analysis

Gene expression data was aggregated by genotype within cell type using AggregateExpression() in the Seurat R package v 4.9.9. Pseudobulk differential expression analysis of the single-cell data was performed on the aggregated count data for each defined cell type with edgeR v 3.40.2 (27) in R v 4.2.0. Patient sex and sequencing pool were added to the statistical model to account for biological variation and batch effects, and a generalized linear model was used to identify differentially expressed genes (DEGs) between each GI disorder group (CD, UC, EoE, and EoD) and controls across eight level one cell types and 29 level two cell types. P-values were adjusted for multiple testing using the Benjamini-Hochberg false discovery rate (FDR), and genes with and FDR < 0.05 were considered significantly differentially expressed.

Functional and pathway analysis for DEGs was carried out using the ToppFun (Transcriptome, ontology, phenotype, proteome, and pharmacome annotations based gene list functional enrichment analysis) tool with default settings from the web-based software ToppGene Suite (http://toppgene.cchmc.org) to identify enriched gene ontology (GO) terms and pathways from databases including KEGG, Reactome, and BioCarta (28). Terms and pathways with a Benjamini-Hochberg FDR < 0.05 and a minimum number of three hits in query list were considered to be significantly enriched.

### Protein-Protein Interaction Networks

Networks of interactions of DEGs were generated in STRING v 12.0 (29) to visualize gene relationships and trends within and between cell types and GI disorders. Networks generated with STRING were imported into Cyotscape v 3.10.0 (30) where node color was used to designate the direction of fold change observed in GI disorders compared to controls, with red corresponding to upregulation and blue corresponding to downregulation. Furthermore, functional terms of interest identified to be enriched in STRING were added to networks.

## Results

### Cell Clustering

A total of 39,622 cells from the 35 individuals in this study passed quality control measures and were mapped to eight level one cell types and 29 level two cell types (**Figure 1**; **Supplementary File 1 Tables A, B, and C**).

**Figure 1.**
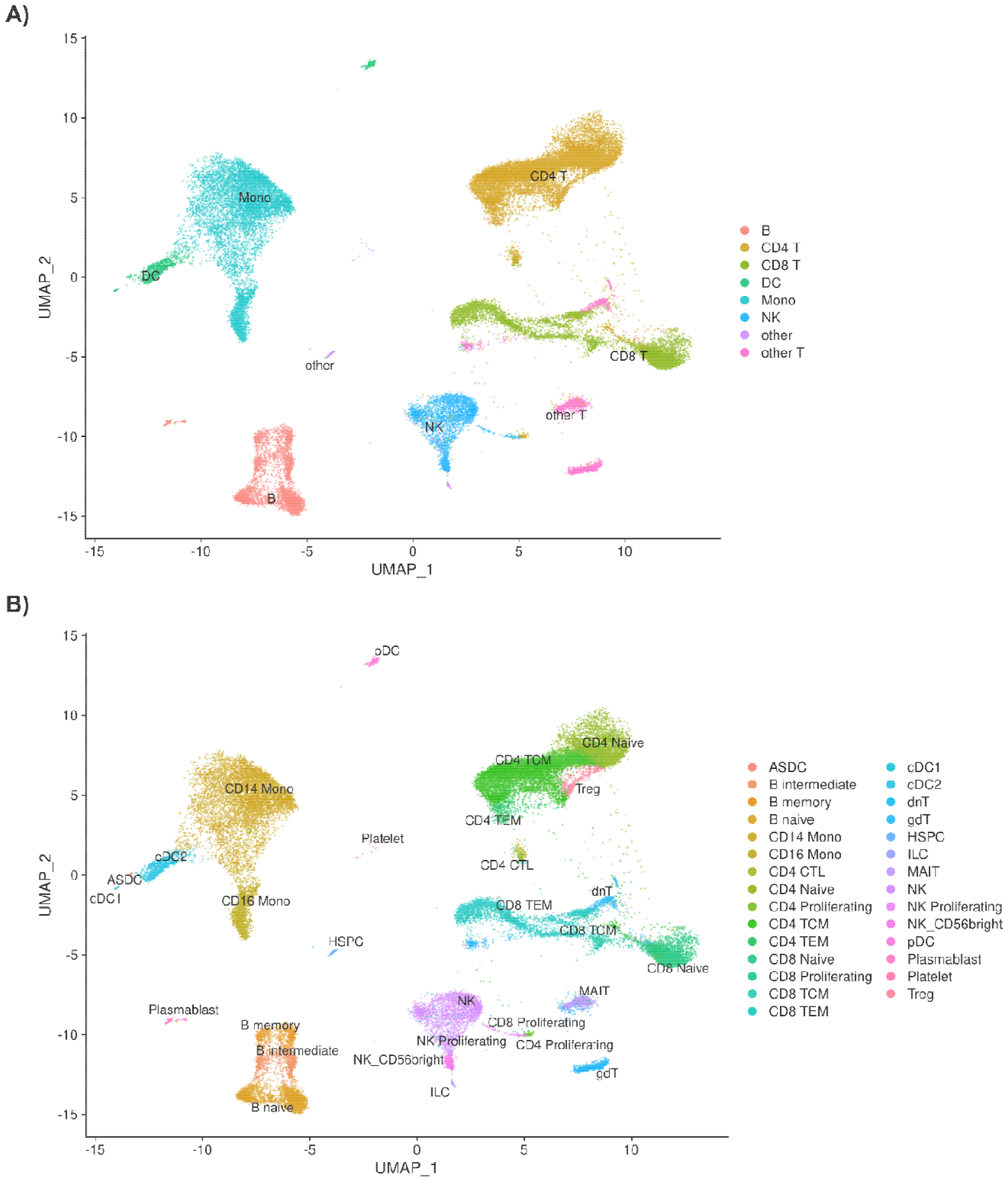
**A)** UMAP of 39,622 cells passing quality control measures sorted into eight level one cell types and into **B)** 29 level two cell types.

### Effect of Gastrointestinal Disorders on the Transcriptome of Immune Cells

Comparison of gene expression between individuals diagnosed with GI disorders (CD, UC, EoD, and EoE) and controls yielded 730 (FDR < 0.05) DEGs (**Figure 2**) across eight level one cell types and 807 (FDR < 0.05) across 25 level two cell types. A list of significant DEGs found across GI disorders within each cell type can be found in **Supplementary File 1 Tables D and E** and volcano plots depicting the results of differential expression analysis for between GI disorders and controls within all level one cell types can be found in **Supplementary File 2 Figures A-H**.

**Figure 2.**
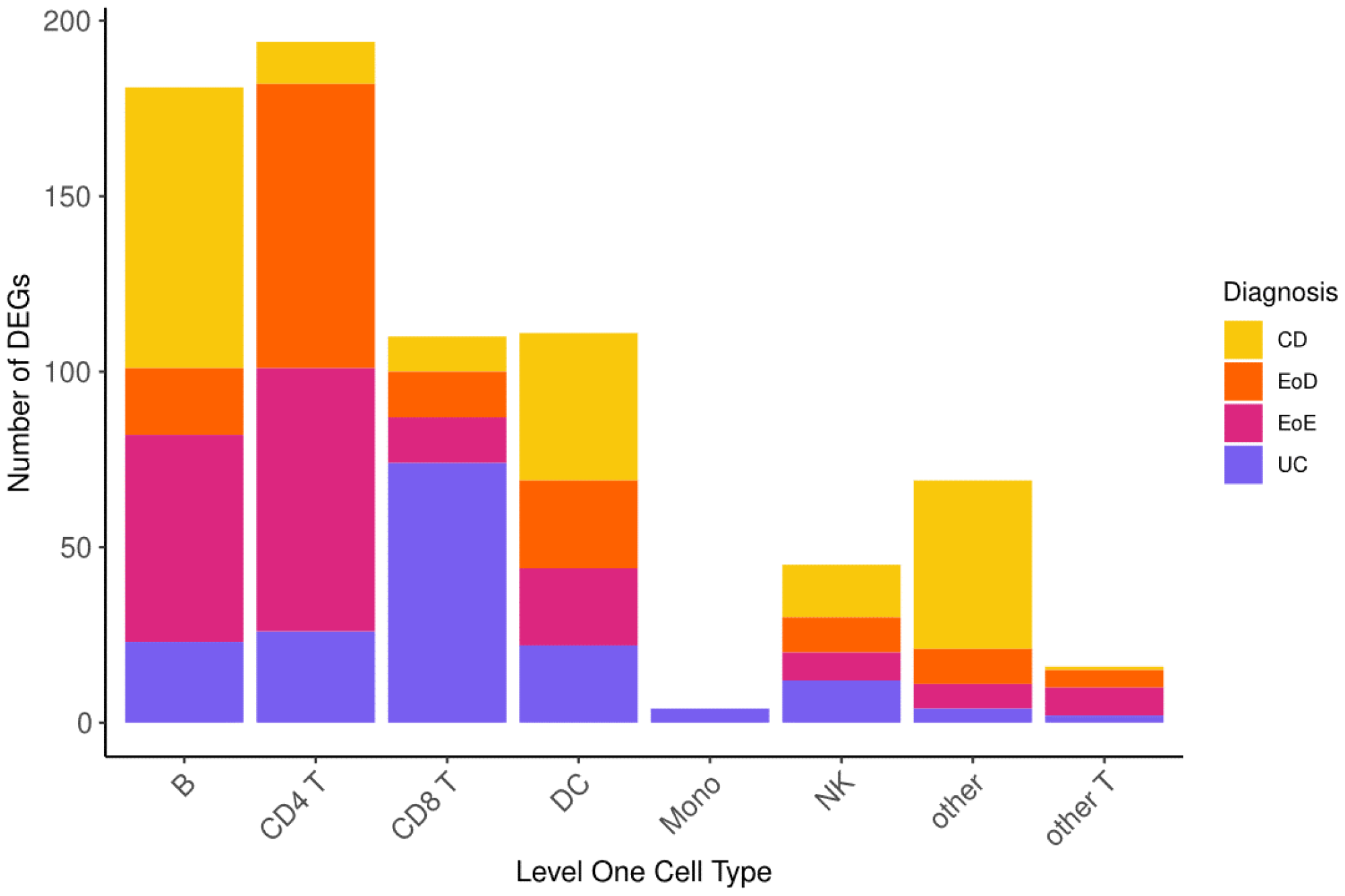
Number of differentially expressed genes (DEGs) across level one cell types and GI disorder subtypes.

Relatively few genes showed common patterns of dysregulation across all GI disorders within cell types: four genes in B cells and DCs, three genes in CD4^+^ and CD8^+^ T cells, and one gene in both NK cells and other T cells (**Supplementary File 1 Table D**). Among these genes with shared patterns of dysregulation were upregulated *MTRNR2L8* in six of the eight level one cell types (B, CD4^+^ T, CD8^+^ T, DC, NK, and other T), downregulated *CPA5* in both CD4^+^ and CD8^+^ T cells, and downregulated *CST3* in CD4^+^ T cells (**Supplementary File 1 Table D**).

Additionally, relatively few genes showed similar patterns of differential expression specific to eosinophilic disorders (EoD and EoE) or IBD (CD and UC) compared to controls. Notable among genes that shared expression patterns among eosinophilic disorders compared to controls was the upregulation of *MTRNR2L1* in four level one cell types (B, CD4^+^ T, CD8^+^ T, and DC), upregulation of several cell cycle associated genes (*MKI67, MELK, CLSPN, CCNA2, TOP2A, MCM5, CDCA8, MYBL2, PCLAF, PCNA, DLGAP5, MCM7, UBE2C, ZWINT, SKA1, CCNB1, CDK1, KIF11, CDKN3*, and *ASPM*) in CD4^+^ T cells (**Figure 3**; **Supplementary File 1 Table D**), and downregulation of *YBX3* in CD8^+^ T cells (**Supplementary File 1 Table D**). In IBD subtypes, *OR11G2* was upregulated in CD4^+^ T and CD8^+^ T cells relative to controls (**Supplementary File 1 Table D**).

**Figure 3.**
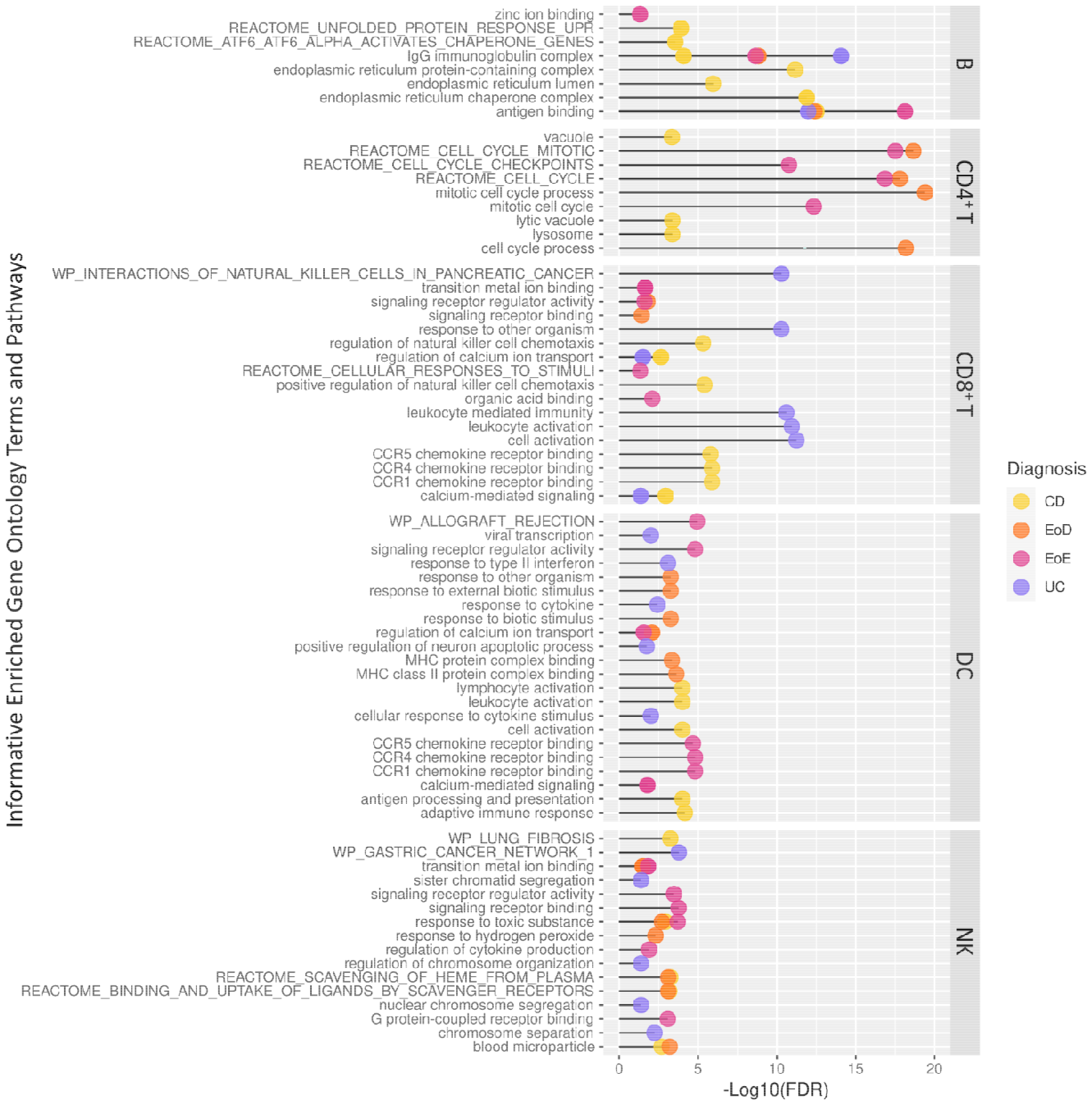
Informative significantly enriched Gene Ontology terms and pathways among differentially expressed genes across level one cell types and GI disorder subtypes identified through ToppGene.

The majority of DEGs did not present with dysregulation in common between GI disorder subtypes; thus, many genes had patterns of dysregulation specific to each GI disorder. Notably, in CD, we observed upregulation of *CC8T* in CD4^+^ T cells, downregulation of *BTN3A2* in CD4^+^ TCM cells, and upregulation of *ETS2* in DCs. In EoD, we observed upregulation of *FOXM1* and *CCR9* in CD4^+^ T cells and downregulation of *GADD45B* and *GADD45G* in CD4^+^ T cells. In EoE, we observed upregulation of *IFNG* in CD4^+^ T cells; upregulation of *IER2* in B and CD8^+^ naïve T cells; upregulation of *EGR1* in three level one cell types (B, CD4^+^ T, and CD8^+^ T cells) and several level two cell types, including naïve CD8^+^ T cells and CD4^+^ TCM cells, and upregulation of both *EGR3* and *EGR4* in B cells. Within UC, we observed downregulation of several genes in CD8^+^ T cells associated with cytotoxicity, such as *CCL5, FGFBP2, GZMA, GZMB, GZMH, IFNG, NKG7*, and *PRF1*; upregulation of *NOG, REG4*, and *AIF1* in CD8^+^ T cells; and upregulation *XIST* in five level one cell types (B, CD4^+^ T, DC, monocytes, and NK) (**Supplementary File 1 Tables D and E**).

A subset of DEGs along with citations of literature supporting their role in GI disorders or in the pathogenesis of inflammation can be found in **Supplementary File 1 Table F**.

### Enrichment of Functional Gene Ontology Terms among DEGs Associated with Gastrointestinal Disorders

Functional analysis of DEGs to identify enriched gene ontology terms and pathways yielded 1037 terms in B cells, 892 terms in CD4^+^ T cells, 760 terms in CD8^+^ T cells, 421 terms in DCs, and 62 terms in NK. To better understand the context of gene dysregulation observed within cell types, and uncover which cellular processes may be affected, functional enrichment analysis of gene ontology terms and pathways was performed. Informative enriched terms for each set of DEGs among GI disorders (CD, UC, EoD, and EoE) within each cell type is shown in **Figure 4**. A full list of significantly enriched GO terms and pathways can be found in **Supplementary File 1 Tables G-K**.

**Figure 4.**
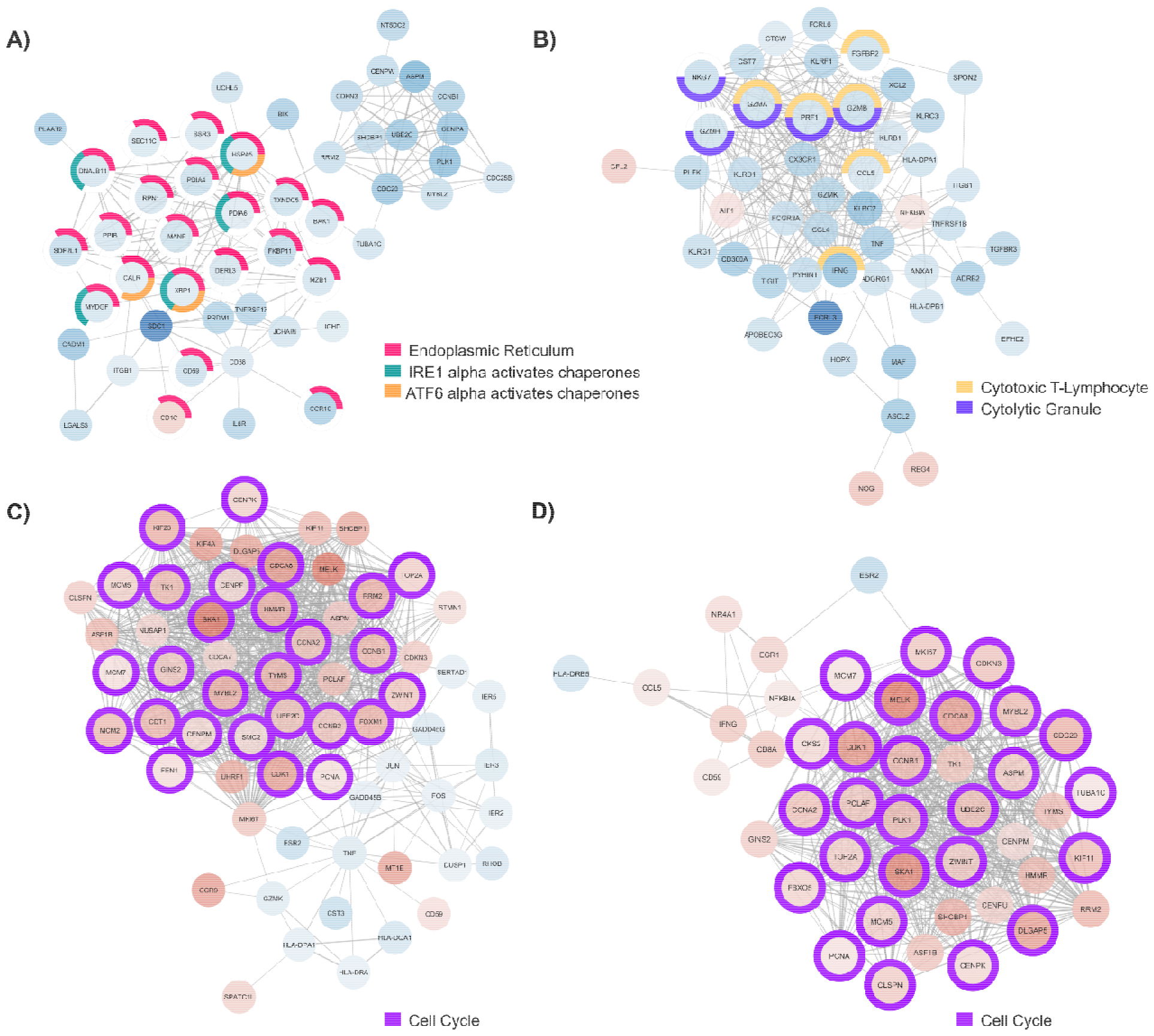
Major component of protein-protein interaction (PPI) network generated with **A)** differentially expressed genes (DEGs) detected in B cells of individuals with CD, **B)** DEGs detected in CD8^+^ T cells from individuals with UC, **C)** DEGs detected in CD4^+^ T cells from individuals with EoD, and **D)** DEGs detected in CD4^+^ T cells from individuals with EoE. Node color indicates the direction of fold change of expression of each gene, with red indicating upregulation of gene expression in patients with GI disorders compared to control and blue indicating downregulation of gene expression in patients with GI disorders compared to control. Node outline color indicates enriched functional annotations detected in STRING.

While there were few enriched GO terms and pathways detected in common among genes dysregulated in CD, UC, EoD, and EoE, there was evidence of altered expression of immunoglobulins in B cells among all GI disorders compared to controls, including antigen binding (GO:0003823) and IgG immunoglobulin complex (GO:0071735) (**Figure 4**; **Supplementary File 1 Table G**). Furthermore, calcium homeostasis-associated terms were enriched among DEGs for each GI disorder, terms included regulation of calcium ion transport (GO:0051924) for DEGs found in CD and UC in CD8^+^ T cells and among DEGs in both EoE and EoD in DCs and regulation of calcium-mediated signaling (GO:0050848) for DEGs detected in CD and UC in CD8^+^ T cells and among DEGs found in EoE within B cells and DCs (**Figure 3**; **Supplementary File 1 Tables G, I, and J**)

Similarities within eosinophilic disorders (EoD and EoE) were also present. GO terms and pathways associated with cell cycle activity in CD4^+^ T cells were enriched among genes differentially expressed in EoD and EoE compared to controls, including Reactome cell cycle mitotic (M5336), Reactome cell cycle (M543) and mitotic cell cycle process (GO:1903047). Other commonly enriched terms among DEGs found in EoD and EoE included transition metal ion binding (GO:0046914) within NK cells and PID AP1 pathway (M167) within CD4^+^ T cells (**Figure 3**; **Supplementary File 1 Tables H and K**)

Specifically, among DEGs in CD, there was an enrichment of GO terms and pathways associated with endoplasmic reticulum function and protein folding in B cells, such as endoplasmic reticulum protein-containing complex (GO:0140534), endoplasmic reticulum chaperone complex (GO:0034663), and Reactome pathways ATF6 alpha activates chaperone genes (M801). Notably, among DEGs in EoD, there was enrichment of terms associated with the p38 MAPK pathway in CD4^+^ T cells, including p38MAPK cascade (GO:0038066) and regulation of p38 MAPK cascade (GO:1900744). Analysis of DEGs within EoE yielded enrichment of zinc ion binding (GO:0008270) within B cells (**Figure 3**; **Supplementary File 1 Tables G and H**).

### Protein-Protein Interaction Networks of DEGs

We generated networks for protein interaction among significant DEGs to identify relationships from curated databases and mined from high-throughput studies and primary literature. A selection of the major components of PPI networks generated in STRING and enhanced with Cytocscape illustrating the relationships of DEGs detected within cell types are shown in **Figure 4**. Additionally, genes associated with enriched gene ontology terms and pathways of interest have been highlighted.

In B cells of individuals with CD, most of the genes associated with endoplasmic reticulum terms are downregulated. Additionally, many of those genes are associated with chaperone functions of the endoplasmic reticulum (**Figure 4A**). We also observed widespread downregulation of DEGs within CD8^+^ T cells among individuals with UC, including markers of cytotoxicity (*CCL5, FGFBP2, GZMA, GZMB, GZMH, IFNG, NKG7*, and *PRF1*) along with the upregulation *NOG, REG4*, and *AIF1* (**Figure 4B**). We further illustrate through PPIs, the similarity of gene dysregulation and dominance of DEGs associated with the cell cycle for both EoE and EoD within CD4^+^ T cells (**Figure 4C and D**), highlighting the similarity of these disorders. However, within these networks we can also see distinct profiles of genes dysregulated between EoE and EoD, including the downregulation of genes associated with p38 MAPK (*GADD45B, GADD45G*, and *DUSP1*) in EoD (**Figure 4C)**.

## Discussion

Aberrant immune signaling due to genetic and environmental factors contributes to the development of GI disorders (31). In this study we focused on the characterization of transcriptomic patterns in PBMCs of pediatric patients with active CD, UC, EoE, and EoD to identify genes and pathways associated with active inflammation and the pathogenesis of each disease. Insights into these transcriptomic phenotypes provide potential indicators of active inflammation and identify genetic markers for improved diagnosis, as well as possible therapeutic targets for treatment.

Genes showing similar patterns of differential expression among all GI disorders (CD, UC, EoE, and EoD) include the upregulation of *MTRNR2L8* within six level one cell types (B, CD4^+^ T, CD8^+^ T, DC, NK, and other T), may be helpful in identifying active inflammation in GI disorders. *MTRNR2L8* is believed to be a marker of cellular stress (32), and upregulation of *MTRNR2L8* has been observed among PBMCs of patients with primary mitochondrial disease (33) and among immune cell types in individuals with aspirin-exacerbated respiratory disease (34).

The common upregulation of *MTRNR2L1* for EoD and EoE was identified in four level one cell types (B, CD4^+^ T, CD8^+^ T, and DC). Similar to *MTRNR2L8, MTRNR2L1* also appears to be upregulated in response to cellular stress (32) and has been found to be upregulated in myeloid cells of patients with autoimmune disorders (35). There was also enrichment of functional terms associated with cell cycle activity in CD4^+^ T cells among DEGs in EoE and EoD compared to controls, prominent among which were Reactome cell cycle mitotic (M5336), Reactome cell cycle (M543) and mitotic cell cycle process (GO:1903047), which may be indicative of dysregulation associated with Th2-mediated inflammation; genes associated with cell cycle progression and proliferation, specifically *MKI76, RRM2*, and *TOP2A*, which were found among upregulated cell cycle genes in EoD and EoE, have also been observed to be upregulated in the epithelium of asthmatics with high levels of Th2 (36).

Functional analysis of DEGs in CD detected in B cells yielded enrichment of terms associated with endoplasmic reticulum function and protein folding. Endoplasmic reticulum stress and the dysregulation of the unfolded protein response have been implicated in the development of IBD, which are essential for the maintenance of homeostasis in both the intestine and immune cells (37, 38). Furthermore, the specific enrichment of Reactome pathway ATF6 alpha activates chaperone genes in CD is supported by evidence of that diminished *ATF6* activity can contribute to intestinal barrier dysfunction in mouse models of IBD (39).

In individual with EoD, upregulation of *CCR9* and downregulation of *GADD45B* and *GADD45G* was found in CD4^+^ T cells of patients compared to controls. *CCR9* has demonstrated to play a key role in Th2-mediated inflammation, with *Ccr9*^-/-^ mice treated with ovalbumin to induce allergic inflammation exhibiting an impaired immune response and diminished recruitment of eosinophils to inflamed tissue (40). Deficiency in *GADD45* expression has been associated with autoimmune disease (41), and the increased expression of *GADD45G* through administration of IL-27, has been shown to attenuate Th2-mediated allergic possibly through the activation of the p38 MAPK pathway (42), a pathway that was found to be enriched among DEGs in CD4^+^ T cells in EoD.

Among identified DEGs in EoE relative to controls was *EGR1* in three level one cell types (B, CD4^+^ T, and CD8^+^ T cells) and several level two cell types, including naïve CD8^+^ T cells and CD4^+^ TCM cells. Among CD4^+^ T cells, *EGR1* is preferentially expressed by Th2 and plays a role in the production of IL-4 cytokines (43), which mediate the allergic inflammatory response (44). Additionally, increased intracellular zinc levels have been shown to upregulate *EGR1* (45). Relevantly, the term zinc ion binding was enriched among DEGs in B cells in EoE. Zinc exposure has been demonstrated to elicit cellular damage (46), induce eosinophilia in mice, and evoke Th2 cytokine production (47). Furthermore, dysregulation of zinc signaling resulting from depletion of zinc within mucosal tissues and the release of zinc from airway discharge has been associated with eosinophilia (48). Together, these data support evidence that zinc homeostasis is critical in regulating inflammatory responses (49) and suggests that zinc homeostasis may be involved the development of eosinophilic disorders.

Several DEGs were found to be dysregulated between UC and controls within CD8^+^ T cells, such as the downregulation of several genes involved in T cell cytotoxicity (*CCL5, FGFBP2, GZMA, GZMB, GZMH, IFNG, NKG7*, and *PRF1*). Previously, it has been shown that while elevated levels of *GZMB* were present in mucosal biopsies of treatment-naïve individuals with CD, there was not upregulation of *GZMB* in treatment-naïve individuals with UC compared to controls, suggesting enhanced CD8^+^ T cytotoxicity in CD but not UC (50). Reduced T-cell cytotoxicity has also been detected in sub-populations of individuals with UC, including those who develop persistent low-grade dysplasia (51). Additionally, widespread upregulation of *XIST* was detected within five level one cell types (B, CD4^+^ T, DC, monocytes, and NK) in UC compared to controls. In mouse models of IBD, *Xist* expression has been demonstrated to be upregulated after inducing colitis, and silencing of *Xist* in these models has shown to reduce colitis-associated symptoms (52). Moreover, previous transcriptome analysis of intestinal mucosa biopsies from individuals with UC have identified *XIST* as a key mediator of inflammation within this disease, as well as a possible therapeutic target (53).

The results of this study hold future implications for clinicians. DEGs that identify active inflammation may be of utility in differentiating patients with active disease and disease in remission without the need for more invasive endoscopy. Additionally, these DEGs show promise of indicating degree of inflammation allowing clinicians to stratify severity of active disease and adjust treatment plans accordingly. Zinc homeostasis dysregulation in eosinophilic disorders provides a potential treatment target in a condition where presently there are few pharmacologic treatment options (54). Patients with EoE are known to have zinc deficiencies associated with elimination diets (55), but further investigation as to how these diets affect overall zinc regulation is needed.

This study demonstrates preliminary evidence for the utility of single-cell RNA-sequencing of patient blood cells to characterize the genomic landscape of pediatric IBD subtypes and eosinophilic disorders. These data indicate both overlapping and distinct DEGs, enriched Gene Ontology terms, and enriched pathways associated the pathogenesis of inflammation among individuals with CD, UC, EoD, and EoE, offering further insight into which genes and pathways may serve as useful markers of disease in the peripheral blood mononuclear cells of patients.

## Data Availability

All data produced in the present study are available upon reasonable request to the authors

## Notes

### Competing Interest Statement

The authors have declared no competing interest.

### Funding Statement

This work was supported, in part, by a generous philanthropic donation made by Emily and Todd Novicoff to the Children's Mercy Kansas City.

### Author Declarations

Ethics committee/IRB of Children's Mercy Hospital gave ethical approval for this work.

### Summary of Updates

Differential gene expression analysis was redone with aggregation performed at the genotype level rather than genotypeXLibrary. The latest version of Seurat has been used. SVAseq was not used in this version, rather sex of patient and pool were added to statistical model to account for unwanted variation.

## References

1. Abraham C, Cho JH. MECHANISMS OF DISEASE Inflammatory Bowel Disease. New Engl J Med. 2009;361(21):2066–78.

2. Silva FAR, Rodrigues BL, Ayrizono MDS, Leal RF. The Immunological Basis of Inflammatory Bowel Disease. Gastroent Res Pract. 2016;2016.

3. Molodecky NA, Soon IS, Rabi DM, Ghali WA, Ferris M, Chernoff G, et al. Increasing incidence and prevalence of the inflammatory bowel diseases with time, based on systematic review. Gastroenterology. 2012;142(1):46–54.

4. Kaplan GG. The global burden of IBD: from 2015 to 2025. Nat Rev Gastroenterol Hepatol. 2015;12(12):720–7.

5. Furuta GT, Katzka DA. Eosinophilic Esophagitis. New Engl J Med. 2015;373(17):1640–8.

6. Bass JA, Friesen CA, Deacy AD, Neilan NA, Bracken JM, Shakhnovich V, et al. Investigation of potential early histologic markers of pediatric inflammatory bowel disease. BMC Gastroenterol. 2015;15:1–7.

7. Jacobs I, Ceulemans M, Wauters L, Breynaert C, Vermeire S, Verstockt B, et al. Role of Eosinophils in Intestinal Inflammation and Fibrosis in Inflammatory Bowel Disease: An Overlooked Villain? Front Immunol. 2021;12:754413.

8. Berg DR, Colombel JF, Ungaro R. The Role of Early Biologic Therapy in Inflammatory Bowel Disease. Inflamm Bowel Dis. 2019;25(12):1896–905.

9. Kelay A, Tullie L, Stanton M. Surgery and paediatric inflammatory bowel disease. Transl Pediatr. 2019;8(5):436–48.

10. Urquhart SA, Quinn KP, Ravi K, Loftus EV, Jr. The Clinical Characteristics and Treatment Outcomes of Concomitant Eosinophilic Esophagitis and Inflammatory Bowel Disease. Crohns Colitis 360. 2021;3(2):otab018.

11. Sawczenko A, Ballinger AB, Savage MO, Sanderson IR. Clinical features affecting final adult height in patients with pediatric-onset Crohn’s disease. Pediatrics. 2006;118(1):124–9.

12. Heuschkel R, Salvestrini C, Beattie RM, Hildebrand H, Walters T, Griffiths A. Guidelines for the management of growth failure in childhood inflammatory bowel disease. Inflamm Bowel Dis. 2008;14(6):839–49.

13. Greenley RN, Hommel KA, Nebel J, Raboin T, Li SH, Simpson P, et al. A meta-analytic review of the psychosocial adjustment of youth with inflammatory bowel disease. J Pediatr Psychol. 2010;35(8):857–69.

14. Cohen NA, Rubin DT. New targets in inflammatory bowel disease therapy: 2021. Curr Opin Gastroenterol. 2021;37(4):357–63.

15. Hart AL, Al-Hassi HO, Rigby RJ, Bell SJ, Emmanuel AV, Knight SC, et al. Characteristics of intestinal dendritic cells in inflammatory bowel diseases. Gastroenterology. 2005;129(1):50–65.

16. Coombes JL, Powrie F. Dendritic cells in intestinal immune regulation. Nat Rev Immunol. 2008;8(6):435–46.

17. Chaudhry A, Rudensky AY. Control of inflammation by integration of environmental cues by regulatory T cells. J Clin Invest. 2013;123(3):939–44.

18. Weaver CT, Elson CO, Fouser LA, Kolls JK. The Th17 pathway and inflammatory diseases of the intestines, lungs, and skin. Annu Rev Pathol. 2013;8:477–512.

19. Ohnmacht C, Marquee R, Presley L, Sawa S, Lochner M, Eberl G. Intestinal microbiota, evolution of the immune systemand the bad reputation of pro-inflammatory immunity. Cellular Microbiology. 2011;13(5):653–9.

20. Mishra A, Hogan SP, Lee JJ, Foster PS, Rothenberg ME. Fundamental signals that regulate eosinophil homing to the gastrointestinal tract. Journal of Clinical Investigation. 1999;103(12):1719–27.

21. Burczynski ME, Peterson RL, Twine NC, Zuberek KA, Brodeur BJ, Casciotti L, et al. Molecular classification of Crohn’s disease and ulcerative colitis patients using transcriptional profiles in peripheral blood mononuclear cells. J Mol Diagn. 2006;8(1):51–61.

22. Martin JC, Chang C, Boschetti G, Ungaro R, Giri M, Grout JA, et al. Single-Cell Analysis of Crohn’s Disease Lesions Identifies a Pathogenic Cellular Module Associated with Resistance to Anti-TNF Therapy. Cell. 2019;178(6):1493–508.

23. Chen P, Zhou GS, Lin JX, Li L, Zeng ZR, Chen MH, et al. Serum Biomarkers for Inflammatory Bowel Disease. Front Med-Lausanne. 2020;7.

24. Kang HM, Subramaniam M, Targ S, Nguyen M, Maliskova L, McCarthy E, et al. Multiplexed droplet single-cell RNA-sequencing using natural genetic variation. Nat Biotechnol. 2018;36(1):89–94.

25. Hao Y, Stuart T, Kowalski MH, Choudhary S, Hoffman P, Hartman A, et al. Dictionary learning for integrative, multimodal and scalable single-cell analysis. Nat Biotechnol. 2024;42(2):293–304.

26. Hao YH, Hao S, Andersen-Nissen E, Mauck WM, Zheng SW, Butler A, et al. Integrated analysis of multimodal single-cell data. Cell. 2021;184(13):3573–87.

27. Robinson MD, McCarthy DJ, Smyth GK. edgeR: a Bioconductor package for differential expression analysis of digital gene expression data. Bioinformatics. 2010;26(1):139–40.

28. Chen J, Bardes EE, Aronow BJ, Jegga AG. ToppGene Suite for gene list enrichment analysis and candidate gene prioritization. Nucleic Acids Res. 2009;37:W305–W11.

29. Szklarczyk D, Kirsch R, Koutrouli M, Nastou K, Mehryary F, Hachilif R, et al. The STRING database in 2023: protein-protein association networks and functional enrichment analyses for any sequenced genome of interest. Nucleic Acids Res. 2023;51(D1):D638-D46.

30. Shannon P, Markiel A, Ozier O, Baliga NS, Wang JT, Ramage D, et al. Cytoscape: A software environment for integrated models of biomolecular interaction networks. Genome Res. 2003;13(11):2498–504.

31. Turpin W, Goethel A, Bedrani L, Croitoru K. Determinants of IBD Heritability: Genes, Bugs, and More. Inflamm Bowel Dis. 2018;24(6):1133–48.

32. Yen K, Lee C, Mehta H, Cohen P. The emerging role of the mitochondrial-derived peptide humanin in stress resistance. J Mol Endocrinol. 2013;50(1):R11–R9.

33. Gordon-Lipkin EM, Banerjee P, Franco JLM, Tarasenko T, Kruk S, Thompson E, et al. Primary oxidative phosphorylation defects lead to perturbations in the human B cell repertoire. Front Immunol. 2023;14:1142634.

34. Bangert C, Villazala-Merino S, Fahrenberger M, Krausgruber T, Bauer WM, Stanek V, et al. Comprehensive Analysis of Nasal Polyps Reveals a More Pronounced Type 2 Transcriptomic Profile of Epithelial Cells and Mast Cells in Aspirin-Exacerbated Respiratory Disease. Frontiers in Immunology. 2022;13.

35. Taft J, Markson M, Legarda D, Patel R, Chan M, Malle L, et al. Human TBK1 deficiency leads to autoinflammation driven by TNF-induced cell death. Cell. 2021;184(17):4447–63.

36. Hachim MY, Elemam NM, Ramakrishnan RK, Salameh L, Olivenstein R, Hachim IY, et al. Derangement of cell cycle markers in peripheral blood mononuclear cells of asthmatic patients as a reliable biomarker for asthma control (vol 11, 11873, 2021). Sci Rep-Uk. 2021;11(1).

37. Cao SS. Endoplasmic Reticulum Stress and Unfolded Protein Response in Inflammatory Bowel Disease. Inflamm Bowel Dis. 2015;21(3):636–44.

38. Li C, Grider JR, Murthy KS, Bohl J, Rivet E, Wieghard N, et al. Endoplasmic Reticulum Stress in Subepithelial Myofibroblasts Increases the TGF-β1 Activity That Regulates Fibrosis in Crohn’s Disease. Inflamm Bowel Dis. 2020;26(6):809–19.

39. Huang SS, Xie Z, Han J, Wang HL, Yang G, Li MY, et al. Protocadherin 20 maintains intestinal barrier function to protect against Crohn’s disease by targeting ATF6. Genome Biol. 2023;24(1).

40. López-Pacheco C, Soldevila G, Du Pont G, Hernández-Pando R, García-Zepeda EA. CCR9 Is a Key Regulator of Early Phases of Allergic Airway Inflammation. Mediat Inflamm. 2016;2016.

41. Schmitz I. Gadd45 Proteins in Immunity. Gadd45 Stress Sensor Genes. 2013;793:51–68.

42. Su XQ, Pan J, Bai FX, Yuan HL, Dong N, Li DD, et al. IL-27 attenuates airway inflammation in a mouse asthma model via the STAT1 and GADD45γ/p38 MAPK pathways. J Transl Med. 2016;14.

43. Lohoff M, Giaisi M, Köhler R, Casper B, Krammer PH, Li-Weber M. Early Growth Response Protein-1 (Egr-1) Is Preferentially Expressed in T Helper Type 2 (Th2) Cells and Is Involved in Acute Transcription of the Th2 Cytokine Interleukin-4. J Biol Chem. 2010;285(3):1643–52.

44. Ricci M, Matucci A, Rossi O. IL-4 as a key factor influencing the development of allergen-specific Th2-like cells in atopic individuals. J Invest Allerg Clin. 1997;7(3):144–50.

45. Barbato JC, Catanescu O, Murray K, DiBello PM, Jacobsen DW. Targeting of metallothionein by L-homocysteine - A novel mechanism for disruption of zinc and redox homeostasis. Arterioscl Throm Vas. 2007;27(1):49–54.

46. Fukui H, Horie M, Endoh S, Kato H, Fujita K, Nishio K, et al. Association of zinc ion release and oxidative stress induced by intratracheal instillation of ZnO nanoparticles to rat lung. Chem Biol Interact. 2012;198(1-3):29–37.

47. Huang KL, Lee YH, Chen HI, Liao HS, Chiang BL, Cheng TJ. Zinc oxide nanoparticles induce eosinophilic airway inflammation in mice. J Hazard Mater. 2015;297:304–12.

48. Suzuki M, Ramezanpour M, Cooksley C, Lee TJ, Jeong B, Kao S, et al. Zinc-depletion associates with tissue eosinophilia and collagen depletion in chronic rhinosinusitis. Rhinology. 2020;58(5):451–9.

49. Devirgiliis C, Zalewski PD, Perozzi G, Murgia C. Zinc fluxes and zinc transporter genes in chronic diseases. Mutat Res-Fund Mol M. 2007;622(1-2):84–93.

50. Jenkins D, Seth R, Kummer JA, Scott BB, Hawkey CJ, Robins RA. Differential levels of granzyme B, regulatory cytokines, and apoptosis in Crohn’s disease and ulcerative colitis at first presentation. J Pathol. 2000;190(2):184–9.

51. Kotsafti A, D’Incà R, Scarpa M, Fassan M, Angriman I, Mescoli C, et al. Weak Cytotoxic T Cells Activation Predicts Low-Grade Dysplasia Persistence in Ulcerative Colitis. Clin Transl Gastroen. 2019;10.

52. Gu D, Cao T, Yi S, Li X, Liu Y. Transcription suppression of GABARAP mediated by lncRNA XIST-EZH2 interaction triggers caspase-11-dependent inflammatory injury in ulcerative colitis. Immunobiology. 2024;229(3):152796.

53. Xu M, Kong Y, Chen N, Peng W, Zi R, Jiang M, et al. Identification of Immune-Related Gene Signature and Prediction of CeRNA Network in Active Ulcerative Colitis. Front Immunol. 2022;13:855645.

54. Tamarit-Sebastian S, Ferrer-Soler FM, Lucendo AJ. Current options and investigational drugs for the treatment of eosinophilic esophagitis. Expert Opin Investig Drugs. 2022;31(2):193–210.

55. Votto M, De Filippo M, Lenti MV, Rossi CM, Di Sabatino A, Marseglia GL, et al. Diet Therapy in Eosinophilic Esophagitis. Focus on a Personalized Approach. Front Pediatr. 2021;9:820192.

