## Supplementary File 2 for "Genomic Insights into Pediatric Intestinal Inflammatory and Eosinophilic Disorders using Single-sell RNA-sequencing"

**Figure A**. Volcano plots showing differentially expressed genes between **A)** CD and control **B)** EoD and control **C)** EoE and control and **D)** UC and control in B cells. Red dots indicate genes upregulated in GI disorders (CD, EoD, EoE, and UC) compared to controls, and blue dots indicate genes downregulated in GI disorders compared to controls.

_­­­­­­­­­­­­­­­_
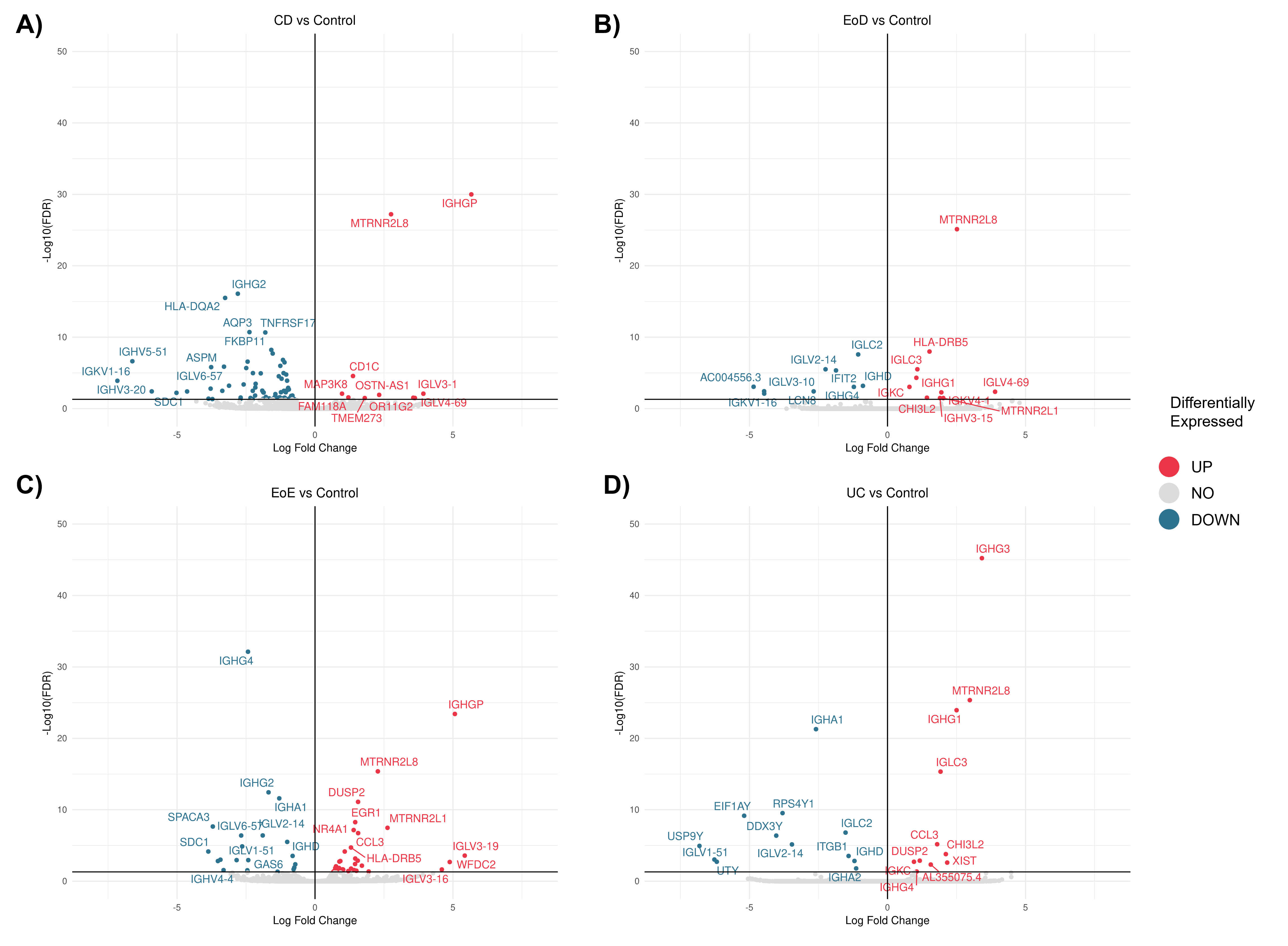

**Figure B**. Volcano plots showing differentially expressed genes between **A)** CD and control **B)** EoD and control **C)** EoE and control and **D)** UC and control in CD4^+^ T cells. Red dots indicate genes upregulated in GI disorders (CD, EoD, EoE, and UC) compared to controls, and blue dots indicate genes downregulated in GI disorders compared to controls.

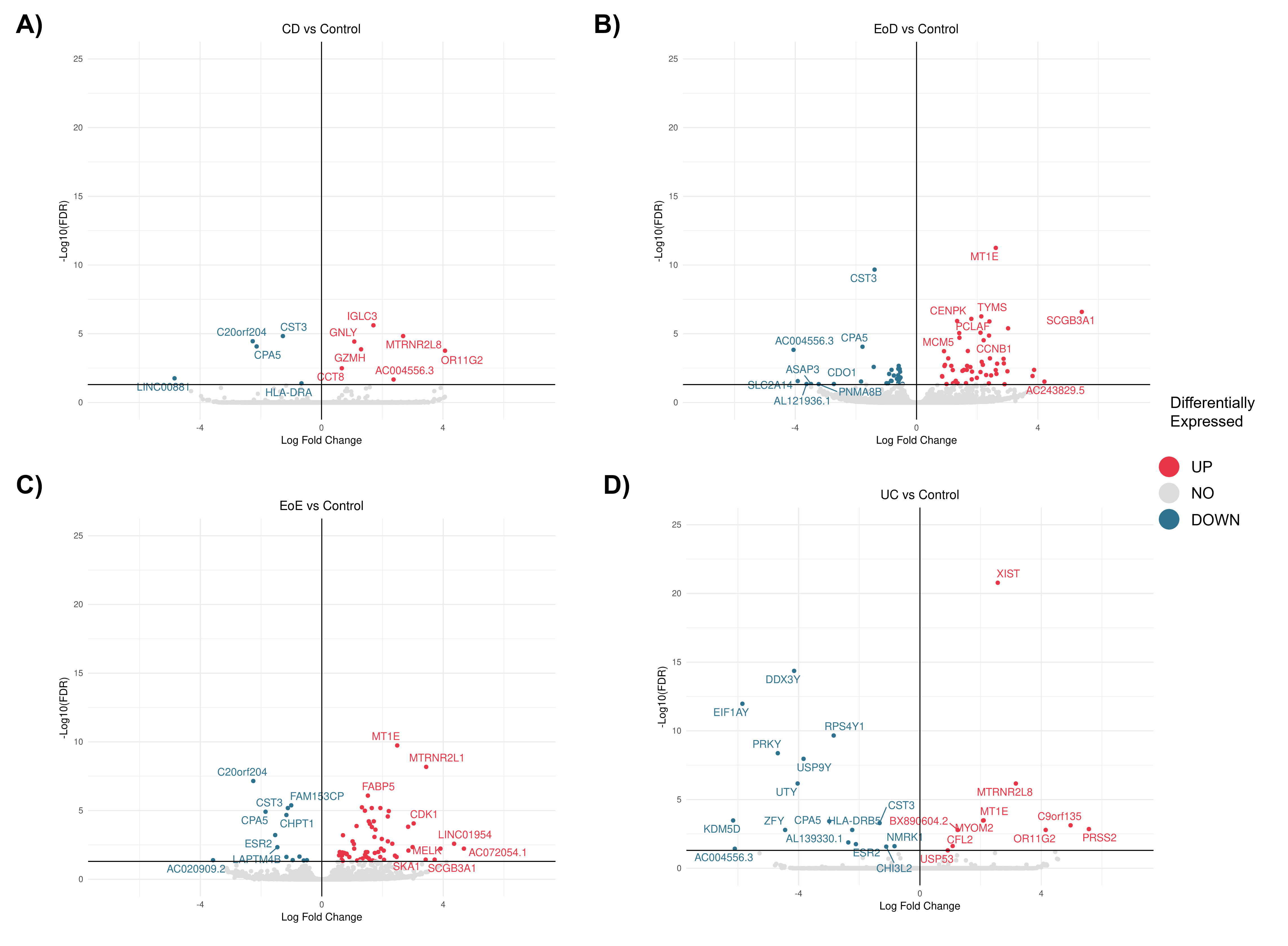

**Figure C**. Volcano plots showing differentially expressed genes between **A)** CD and control **B)** EoD and control **C)** EoE and control and **D)** UC and control in CD8^+^ T cells. Red dots indicate genes upregulated in GI disorders (CD, EoD, EoE, and UC) compared to controls, and blue dots indicate genes downregulated in GI disorders compared to controls.

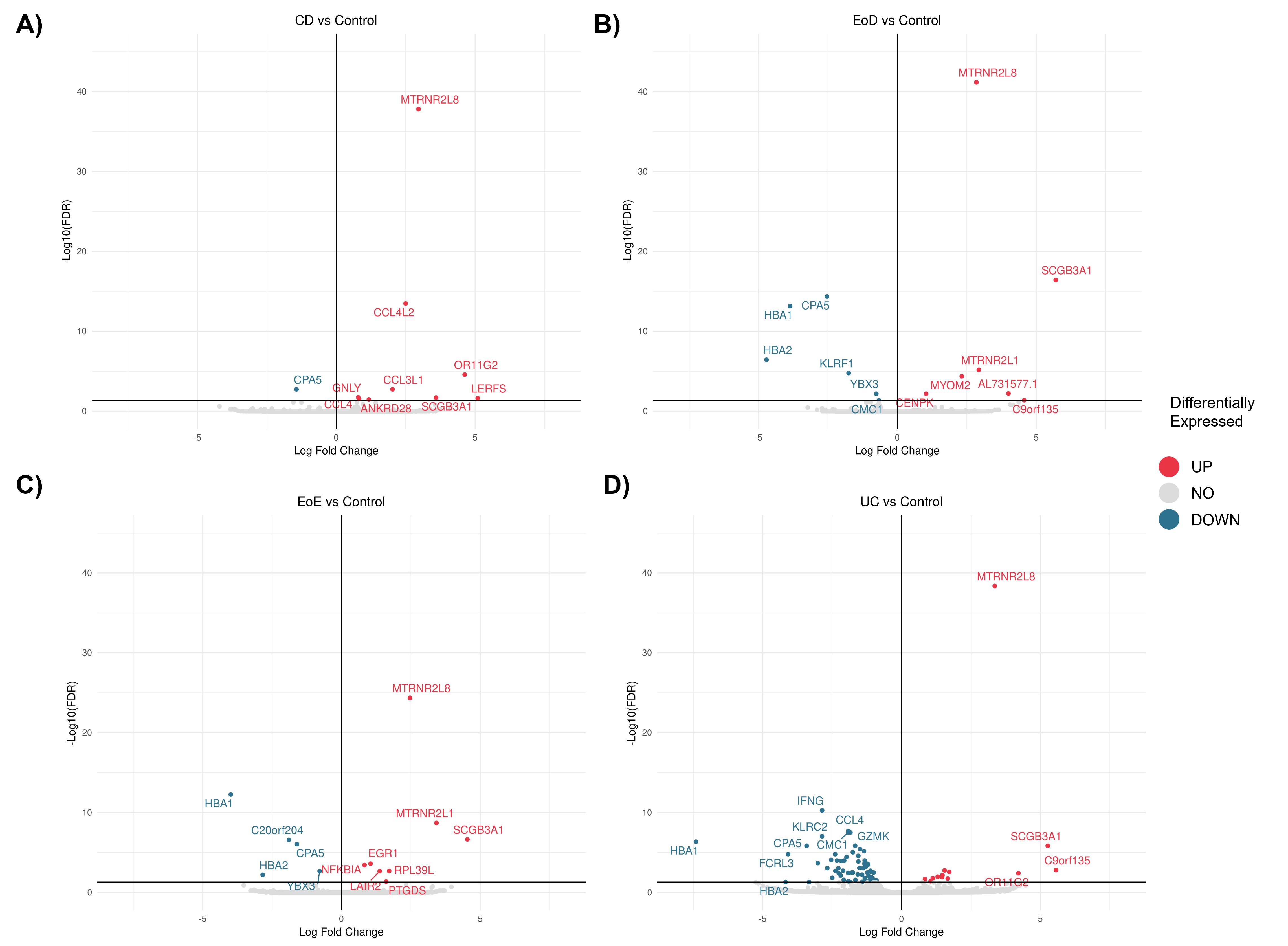

**Figure D**. Volcano plots showing differentially expressed genes between **A)** CD and control **B)** EoD and control **C)** EoE and control and **D)** UC and control in dendritic cells. Red dots indicate genes upregulated in GI disorders (CD, EoD, EoE, and UC) compared to controls, and blue dots indicate genes downregulated in GI disorders compared to controls.

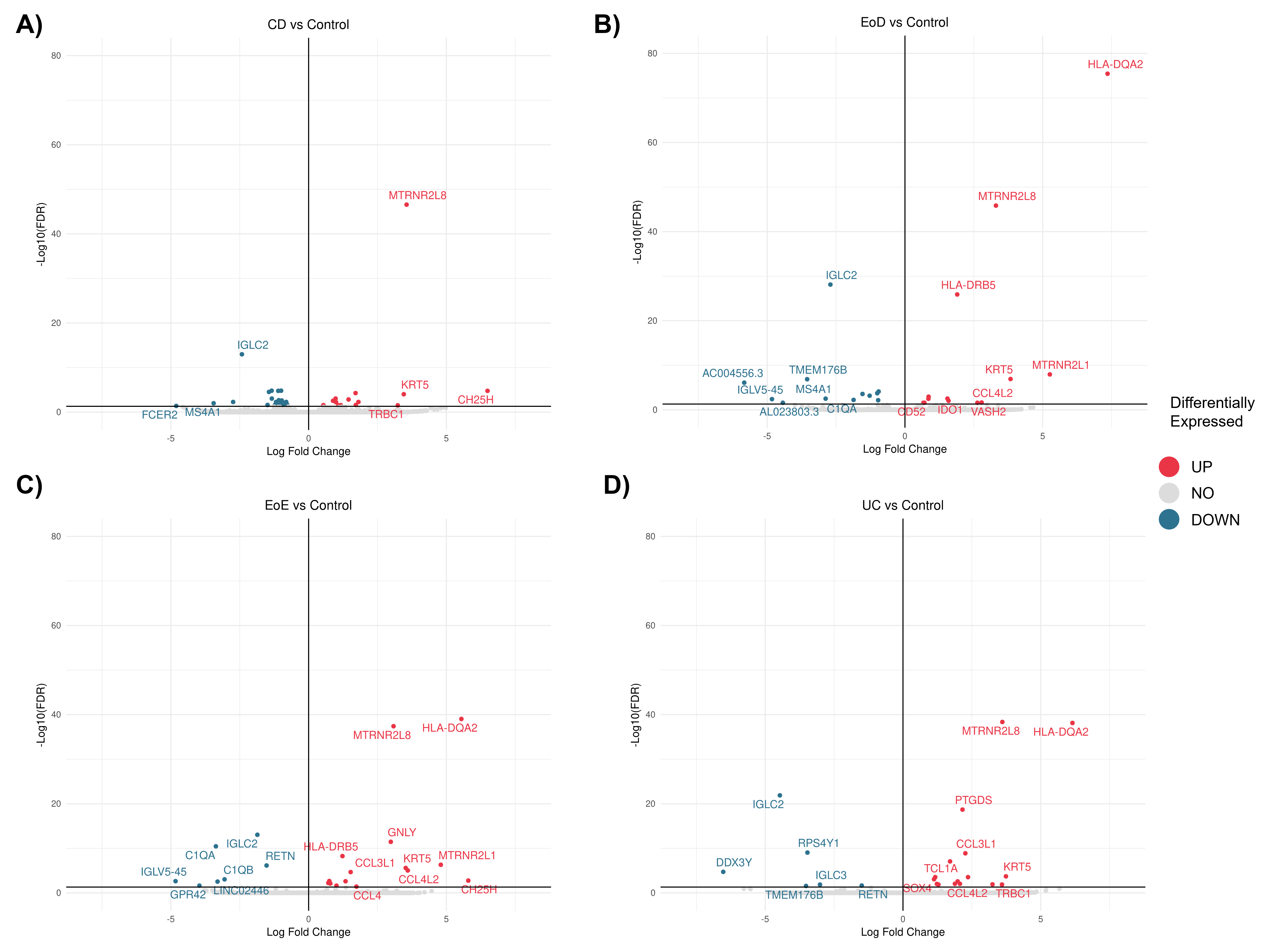

**Figure E**. Volcano plots showing differentially expressed genes between **A)** CD and control **B)** EoD and control **C)** EoE and control and **D)** UC and control in monocytes. Red dots indicate genes upregulated in GI disorders (CD, EoD, EoE, and UC) compared to controls, and blue dots indicate genes downregulated in GI disorders compared to controls.

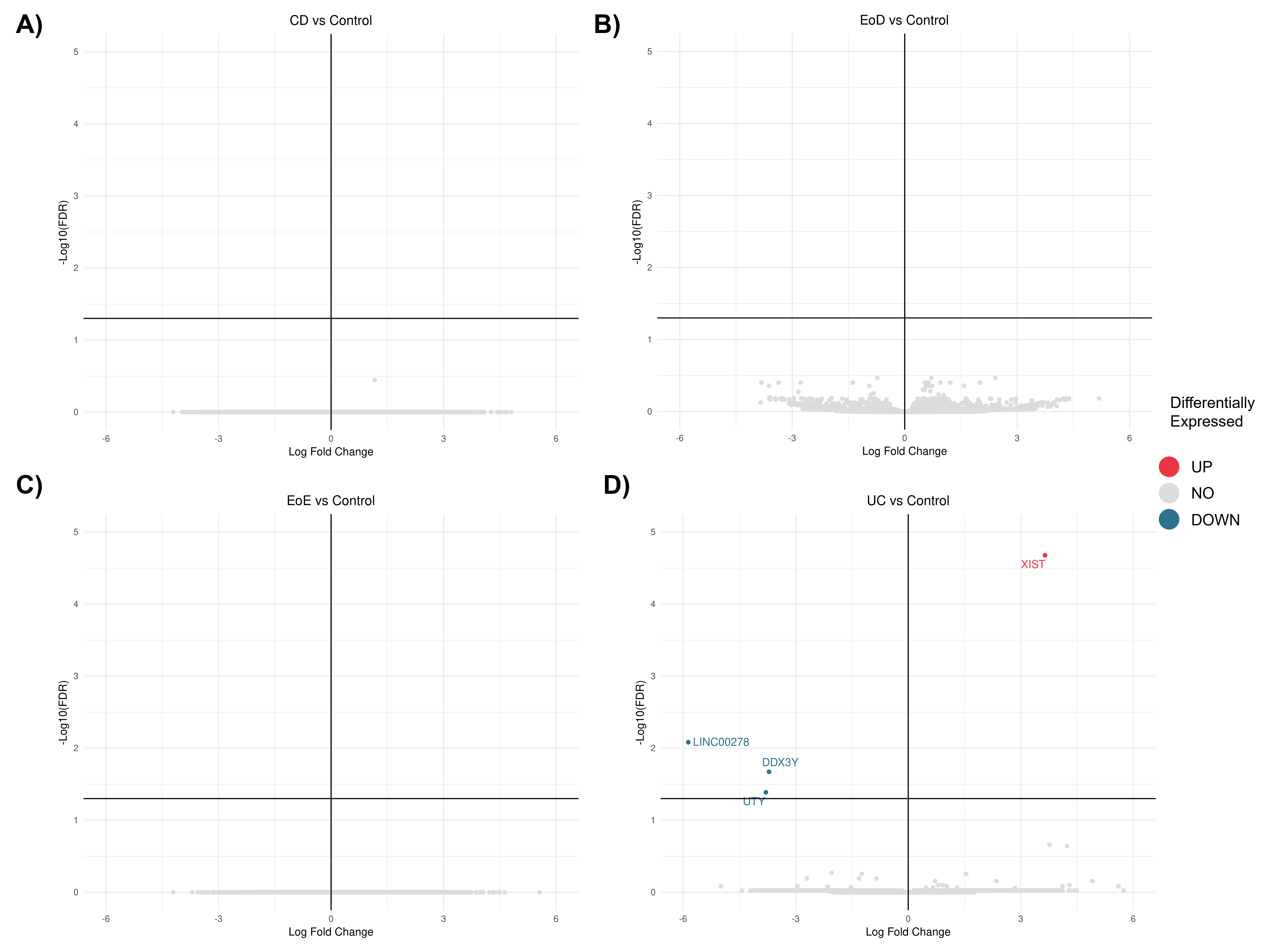

**Figure F**. Volcano plots showing differentially expressed genes between **A)** CD and control **B)** EoD and control **C)** EoE and control and **D)** UC and control in natural killer cells. Red dots indicate genes upregulated in GI disorders (CD, EoD, EoE, and UC) compared to controls, and blue dots indicate genes downregulated in GI disorders compared to controls.

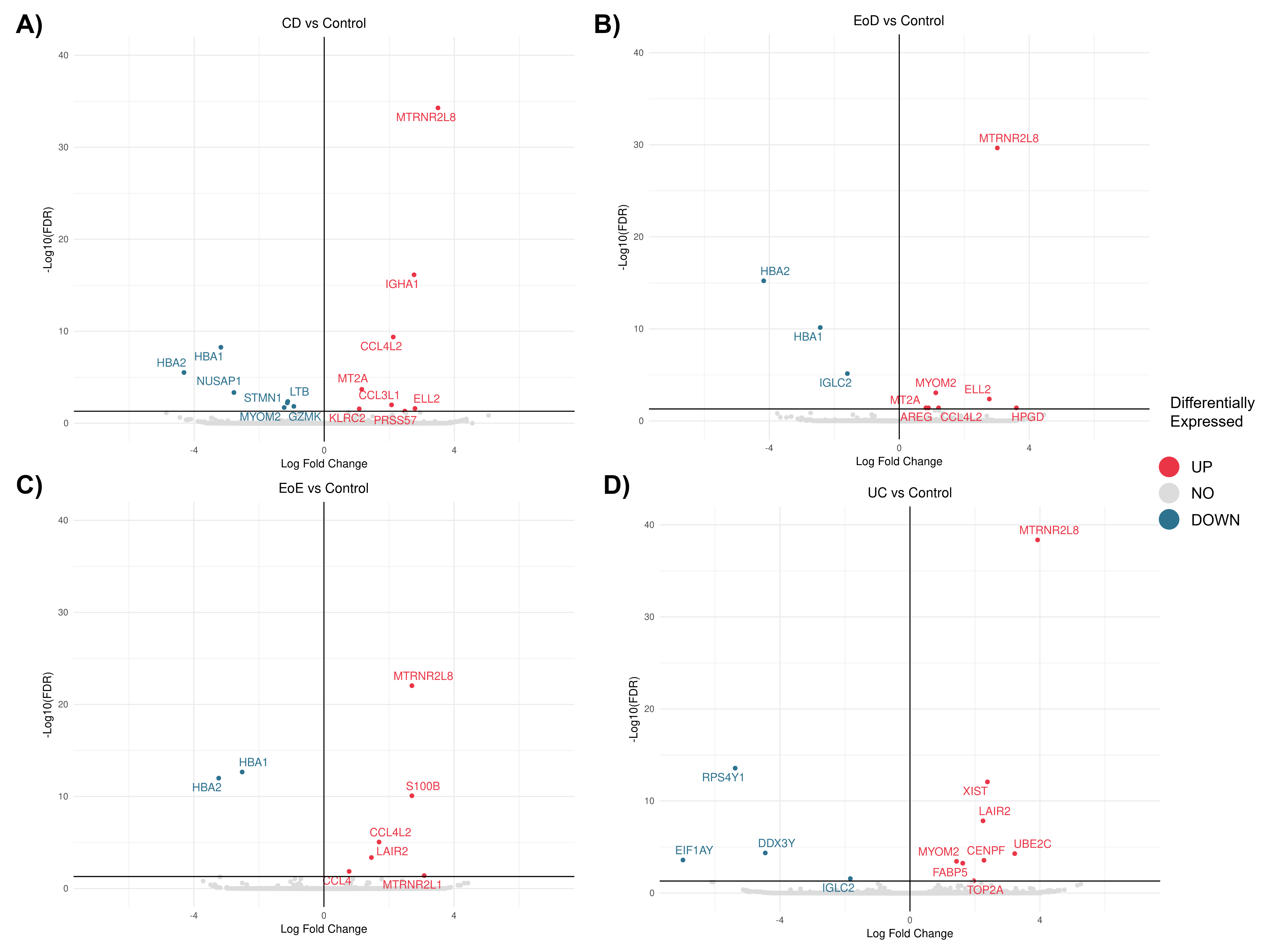

**Figure G**. Volcano plots showing differentially expressed genes between **A)** CD and control **B)** EoD and control **C)** EoE and control and **D)** UC and control in other T cells. Red dots indicate genes upregulated in GI disorders (CD, EoD, EoE, and UC) compared to controls, and blue dots indicate genes downregulated in GI disorders compared to controls.

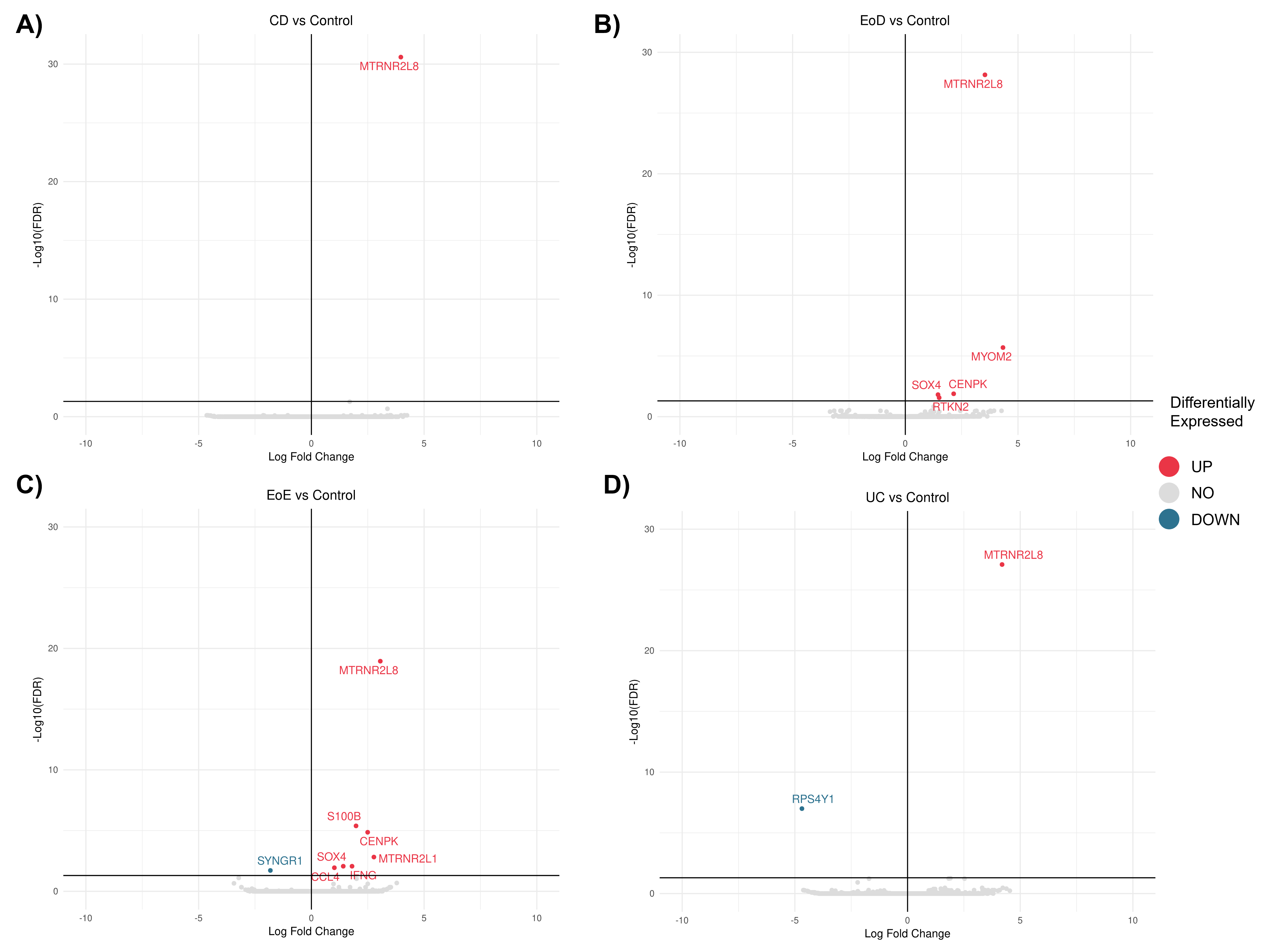

**Figure H**. Volcano plots showing differentially expressed genes between **A)** CD and control **B)** EoD and control **C)** EoE and control and **D)** UC and control in other cells. Red dots indicate genes upregulated in GI disorders (CD, EoD, EoE, and UC) compared to controls, and blue dots indicate genes downregulated in GI disorders compared to controls.

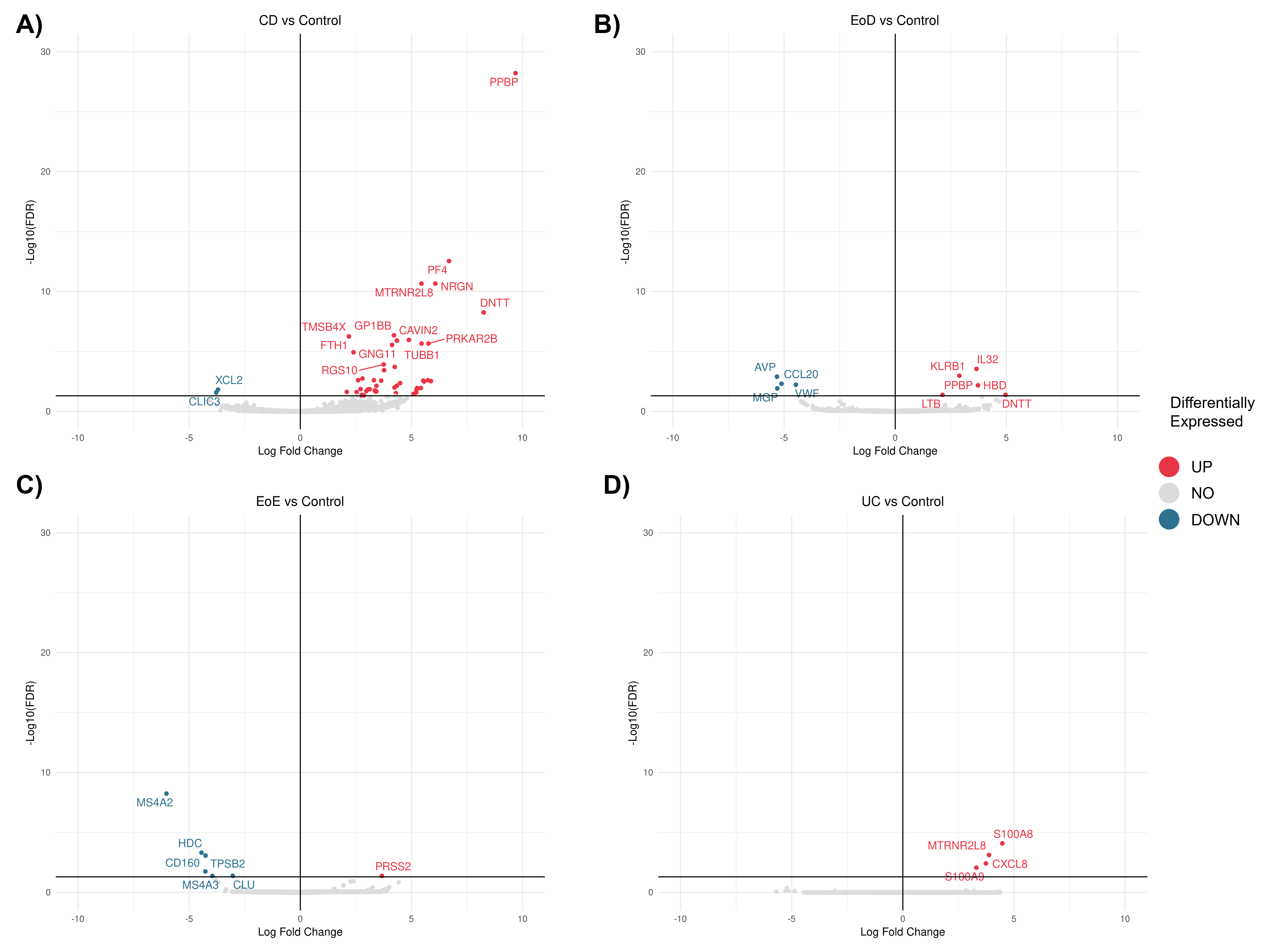
